# Understanding and Predicting Polycystic Ovary Syndrome through Shared Genetic Architecture with Testosterone, SHBG, and Inflammatory Markers

**DOI:** 10.1101/2023.10.17.23297115

**Authors:** Lillian Kay Petersen, Garyk Brixi, Jun Li, Jie Hu, Zicheng Wang, Xikun Han, Anat Yaskolka Meir, Jaakko Tyrmi, Shruthi Mahalingaiah, Terhi Piltonen, Liming Liang

## Abstract

Polycystic ovary syndrome (PCOS) is a common hormonal disorder that affects one out of eight women and has high metabolic and psychological comorbidities. PCOS is thought to be associated with obesity, hormonal dysregulation, and systemic low-grade inflammation, but the underlying mechanisms remain unclear. Here we study the genetic relationship between PCOS and obesity, testosterone, sex hormone binding globulin (SHBG), and a wide-range of inflammatory markers. First, we created a large meta-analysis of PCOS (7,747 PCOS cases and 498,227 controls) and identified four novel genetic loci associated with PCOS. These novel loci have been previously associated with gene expression in multiple PCOS-relevant tissues including the thyroid and ovary. We then further incorporated GWASs for obesity (n=681,275), SHBG (n=190,366), testosterone (n=176,687), and 138 inflammatory biomarkers (average n=30,000). Using Mendelian randomization methods, we replicated genetic causal relationships from obesity and SHBG to PCOS. We identified significant genetic correlations between PCOS and eleven inflammatory biomarkers, including novel and strong correlations with death receptor 5 (LDSC rg = 0.54, FDR = 0.043), among others. Although no statistically significant causal relationship was observed between inflammatory markers and PCOS, 31 inflammatory biomarkers showed significant causal effects on SHBG or testosterone, supporting a potentially etiological role of chronic inflammation in influencing sex hormone levels. Finally, we show that combining the polygenic risk scores of PCOS and PCOS-related traits improves genetic prediction of PCOS cases in the UK Biobank and MGB Biobank, as compared to using only the risk score of PCOS. Together, these results support the theory that immune responses are altered in PCOS patients and that chronic inflammation may play a role in testosterone dysregulation.

## 1 Introduction

Polycystic ovary syndrome (PCOS) is a complex endocrine disorder that is estimated to affect between 5-20% of women of reproductive age [1]. PCOS is mainly diagnosed using the Rotterdam criteria, which requires the presence of two out of the following three symptoms: biochemical or clinical hyperandrogenism, irregular menstruation or anovulation, and polycystic ovarian morphology [2]. Women with PCOS report lowered work ability and quality of life [3, 4, 5]. PCOS has been related to comorbidities including metabolic, reproductive and psychological disorders, obesity, diabetes, dyslipidemia, metabolic syndrome, obstructive sleep apnea, cardiovascular disease, subfertility, endometrial cancer, depression, anxiety, and eating disorders [6].

The pathogenesis of PCOS includes both genetic and environmental factors, but the specific mechanisms remain unclear [7]. PCOS is polygenic and highly heritable (heritability *>*70% based on twin studies) [8]. Genome-wide association studies (GWAS) have identified 22 genetic loci associated with PCOS, but the proportion of heritability explained remains low with a limited number of functional studies [9, 10, 11]. Hyperandrogenism and PCOS pathogenesis have been linked to factors such as weight gain, obesity, and insulin resistance; chronic low-grade inflammation [12]; and low sex hormone binding globulin (SHBG) levels [13]. Previous studies have found significant genetic correlations between PCOS, BMI, and waist-to-hip ratio (WHR) [14], and a potential causal role of BMI, type 2 diabetes, SHBG, and other traits in the development of PCOS based on previous Mendelian randomization (MR) studies [15]. PCOS patients often have elevated inflammation markers compared with age- and BMI-matched controls [16, 17, 18], and anti-inflammatory therapy can reverse PCOS-like traits in animal models [7]. This supports a potential role for altered immune response in PCOS.

Here we investigate the genetic relationship between PCOS, obesity, testosterone, and SHBG, and how this relationship is connected with and potentially mediated by inflammation. Figure 1 shows an overview of our study design.

**Figure 1:**
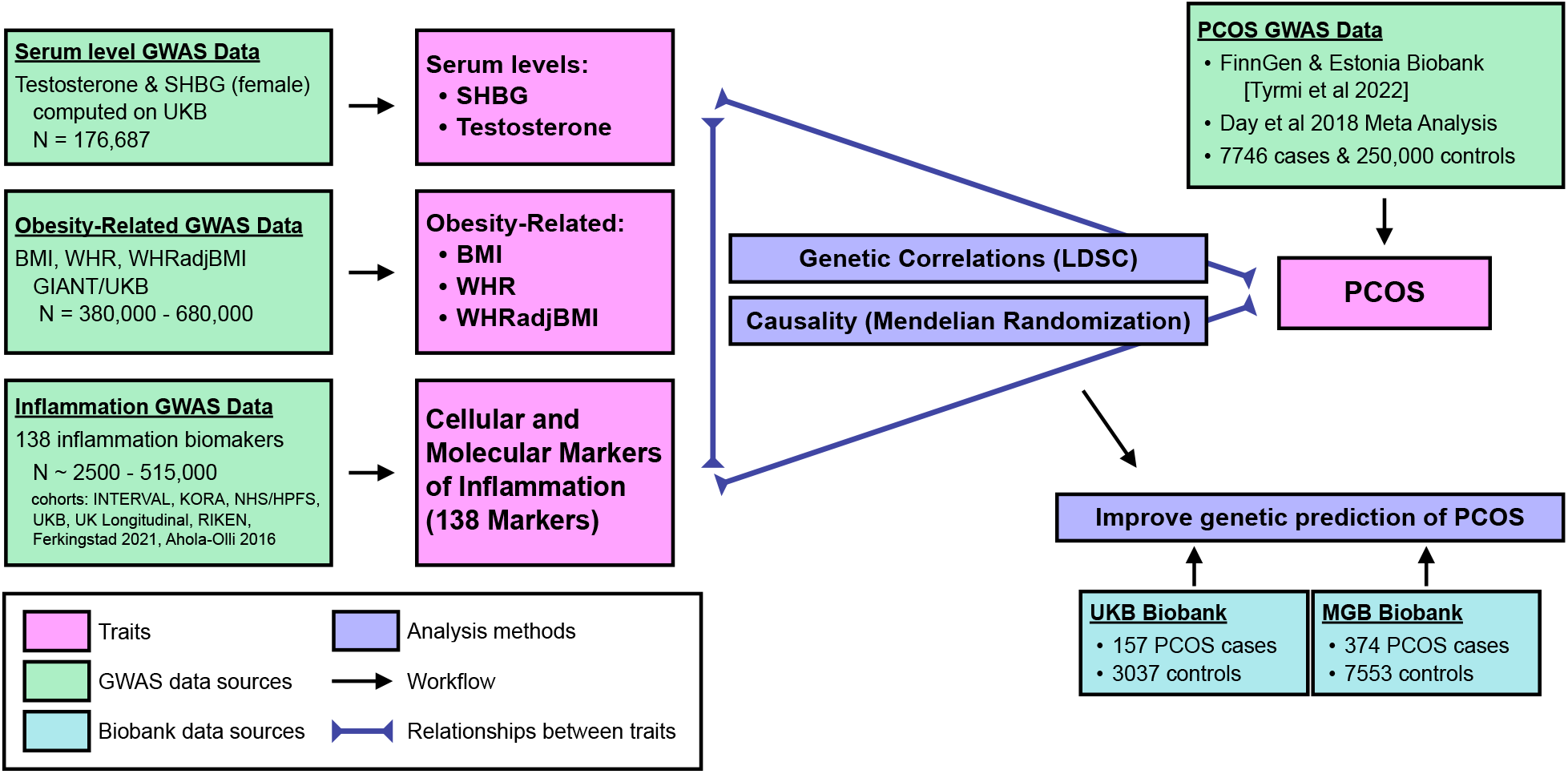
Flowchart depicting data sources and analysis strategies.

## 2 Results

### 2.1 PCOS Meta-analysis

We first combined Tyrmi et al. [9] and Day et al. [10] summary statistics to create a large meta-analysis of PCOS. The meta-analysis identified 26 genome-wide significant (*p <* 5 *×* 10^*−*8^) linkage disequilibrium (LD) independent loci with significant associations with PCOS. Four of thwese loci have not been previously reported to be associated with PCOS in the literature: rs61030588, rs11234902, rs56738967, rs78378222. Details about the lead variants are shown in table 1, and the full meta-analysis results are shown in figure 2.

**Table 1:** Loci that reach genome-wide significance for PCOS in the meta-analysis and (top) are in LD with loci previously reported for PCOS; or (bottom) are not in LD with loci previously reported for PCOS.

| CHR:BP Ref/Effect | RSID | Beta | Effect Allele Freq | p-value | Nearest Gene |
| --- | --- | --- | --- | --- | --- |
| **Replicated PCOS Loci** |  |  |  |  |  |
| 2: 43561780 G/A | rs7563201 | -0.108 | 0.521 | $5.93 \times 10^{-8}$ | *THADA* |
| 2: 213387900 C/T | rs7564590 | 0.144 | 0.645 | $1.90 \times 10^{-12}$ | *ERBB4* |
| 5: 16836005 C/T | rs9312937 | -0.125 | 0.457 | $1.90 \times 10^{-12}$ | *MYO10* |
| 8: 11621450 G/A | rs2740433 | -0.140 | 0.344 | $1.91 \times 10^{-11}$ | *GATA4* |
| 9: 126637668 G/T | rs7028482 | -0.304 | 0.065 | $1.49 \times 10^{-14}$ | *DENND1A* |
| 11: 30226356 C/T | rs11031005 | -0.202 | 0.142 | $8.31 \times 10^{-14}$ | *FSHB* |
| 11: 113949232 C/T | rs1784692 | 0.203 | 0.138 | $3.69 \times 10^{-12}$ | *ZBTB16* |
| 12: 75978358 G/A | rs1148006 | -0.117 | 0.705 | $4.70 \times 10^{-8}$ | *KRR1* |
| 22: 29098376 G/A | rs182075939 | -0.523 | 0.046 | $1.93 \times 10^{-16}$ | *CHEK2* |
| **New PCOS Loci** |  |  |  |  |  |
| 2: 47995854 G/A | rs61030588 | -0.125 | 0.283 | $3.42 \times 10^{-8}$ | *MSH6* |
| 11: 86712340 G/A | rs11234902 | -0.141 | 0.762 | $7.42 \times 10^{-10}$ | *FZD4-DT* |
| 16: 79740541 G/C | rs56738967 | 0.117 | 0.695 | $2.46 \times 10^{-8}$ | *MAF* |
| 17: 7571752 G/T | rs78378222 | -0.405 | 0.021 | $4.47 \times 10^{-8}$ | *TP53* |

**Figure 2:**
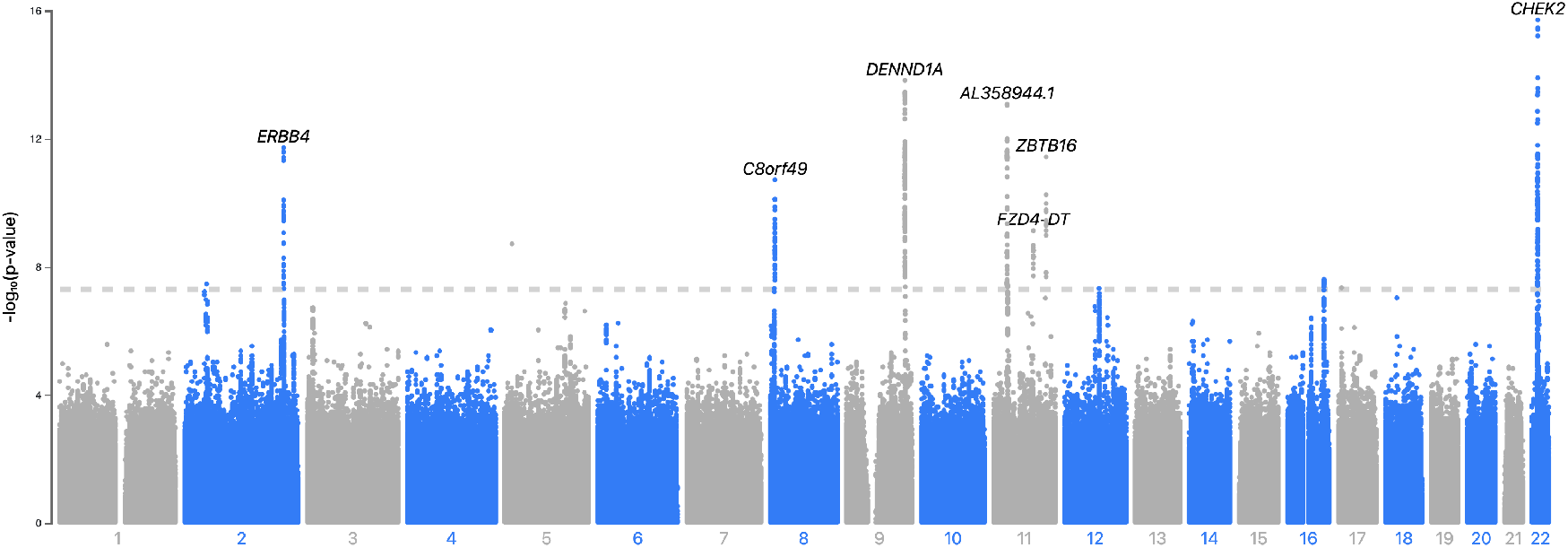
Manhattan plot of PCOS meta-analysis

**Figure 3:**
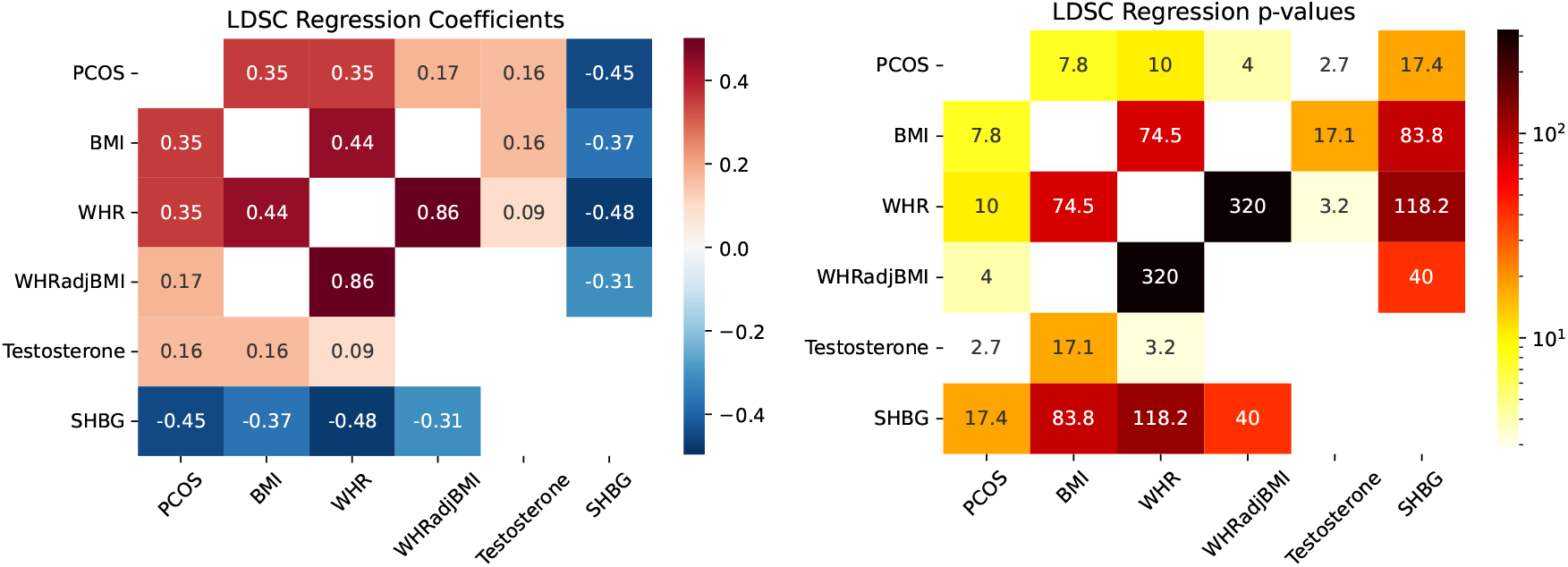
Genetic correlations and p-values (-log10), computed via LDSC, between the non-inflammatory traits in our study. Values that did not meet FDR *<* 0.05 are not shown.

All of the identified novel loci for PCOS were significant expression quantitative trait (eQTL) loci in multiple tissues [19]. Rs61030588 on chromosome 2 is within the gene *MSH6* and is a known eQTL in multiple tissues including the thyroid and ovaries. *MSH6* is a gene involved in DNA repair, which has previously been implicated in menopause age and ovarian aging [20]. The novel PCOS locus rs11234902 on chromosome 11 is a significant thyroid tissue eQTL for *RP11-736K20*.*6*, an RNA gene. The novel locus on chromosome 16, rs56738967, is a significant eQTL for *MAFTRR* in a large number of tissues including the thyroid and ovary. *MAFTRR* is a lncRNA involved in gene regulation. The new locus on chromosome 17 is on an intronic region of *TP53*, a tumor suppressor gene. This variant has been reported to increase risk of uterine fibroids, gliomas, and lean mass, while other variants on *TP53* are inked to levels of SHBG and testosterone [21, 22, 23]. The rs78378222 locus is a significant eQTL for *TP53* in several tissues including adipose tissue. The full tables of eQTL hits for novel PCOS loci lead variants are included in the supplementary file 1.

### 2.2 Genetic Correlation

#### 2.2.1 Genetic correlations between PCOS, obesity, testosterone, and SHBG

We curated GWAS summary statistics for body mass index (BMI) and female-specific GWAS for SHBG, testosterone, waist-to-hip ratio (WHR), and female-specific waist-hip ratio adjusted for BMI (WHRadjBMI), and computed the genetic correlations between these traits with PCOS using LDSC.

The genetic correlations between these PCOS-related traits are shown in table 3. PCOS has strong genetic correlations with SHBG (r_*g*_=0.45), BMI (r_*g*_=0.35), and WHR (r_*g*_=0.35), and has weaker correlations with WHRadjBMI (r_*g*_=0.17) and testosterone (r_*g*_=0.16). SHBG is inversely correlated with all of obesity-related traits especially WHR. We did not find significant correlations between testosterone and SHBG.

#### 2.2.2 Genetic correlations of inflammatory biomarkers with PCOS, obesity, testosterone, and SHBG

We created meta-analyses of 138 inflammatory biomarkers, including immune cell counts and biomarker serum levels. The sample sizes of these biomarkers ranged between 2,538 and 505,690 individuals, and SNP-based heritability ranged from 0 to 0.5. The full table describing these inflammatory biomarkers are shown in the supplementary file 2.

We first examined genetic correlations between inflammatory biomarkers and PCOS, SHBG and testosterone. For inflammatory biomarkers with significant genetic correlations with at least one of the three traits (FDR *<* 0.05), we further examined their genetic correlations with obesity traits (as shown in figure 4, with the values included in supplementary file 3).

**Figure 4:**
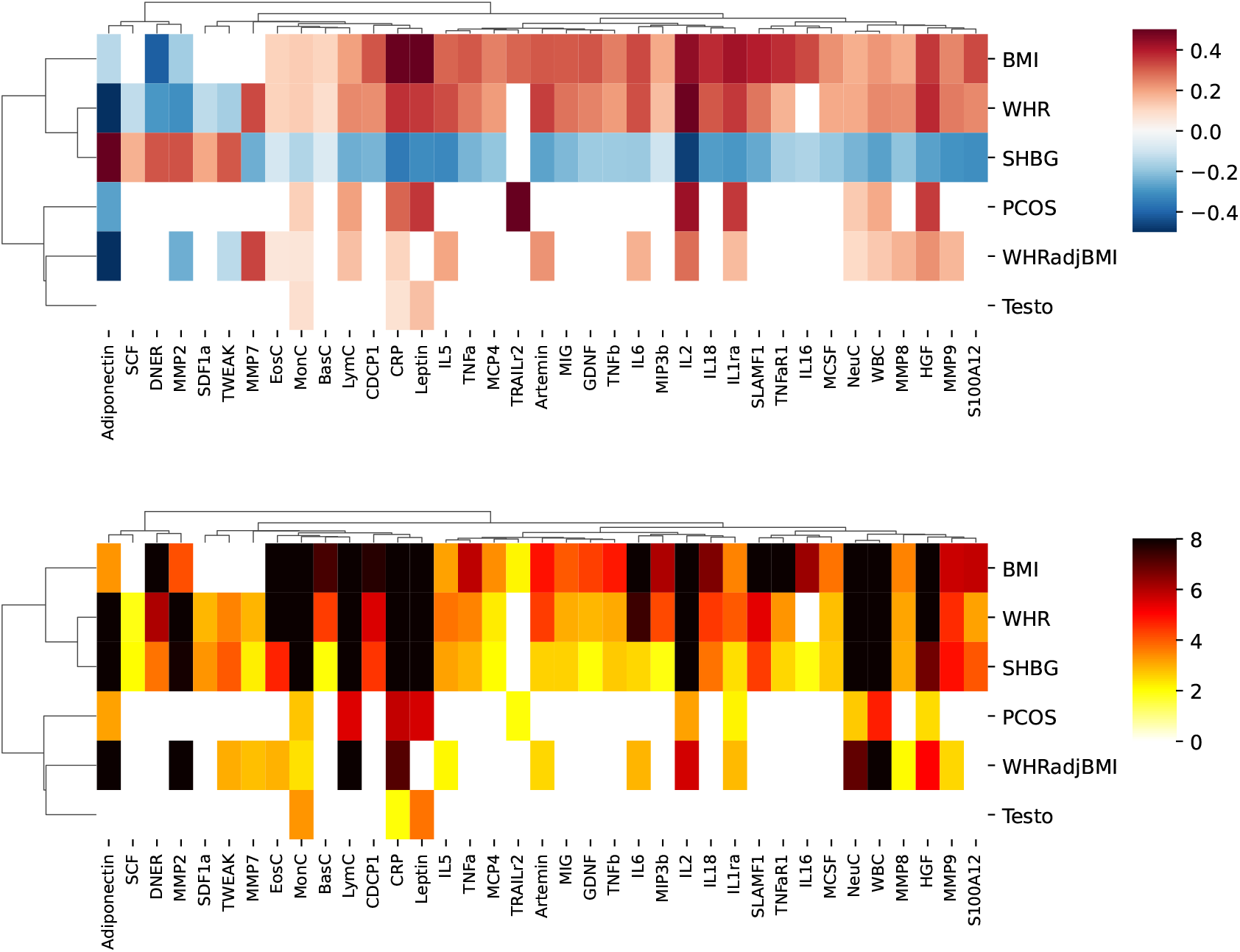
Genetic correlations (top) and -log10 p-value (bottom) computed via LDSC between the traits PCOS, BMI, WHR, WHRadjBMI, SHBG, testosterone and the inflammation markers. Correlations and p-values shown are FDR *<* 0.05. Rows and columns are clustered by genetic correlation using hierarchical clustering, and we reversed the correlation direction of SHBG before clustering.

PCOS showed significant genetic correlation with TRAILr2, IL2, leptin, IL1ra, HGF, CRP, adiponectin, as well as circulating counts of total white blood cells (WBC), lymphocytes (LymC), neutrophils (NeuC), and monocytes (MonC). Most of these inflammatory biomarkers that correlated with PCOS showed significant genetic correlation of similar magnitude with BMI, and WHR, or negative SHBG. The exception was TRAILr2, which is only related to BMI but not WHR.

SHBG is significantly genetically correlated with 36 inflammation markers, all of which were also significantly correlated to an obesity trait in the opposite directions. Testosterone only showed significant genetic correlations with three inflammatory markers, namely with leptin, CRP, and monocyte count.

Hierarchical clustering on the inflammatory LDSC correlations suggests that WHR, BMI, and low SHBG share a similar inflammatory profile, while PCOS has a inflammatory profile closer to WHRadjBMI and testosterone.

#### 2.2.3 Causal relationships between PCOS, obesity, testosterone, and SHBG

To investigate potential causal relationships, we conducted bi-directional MR analysis between PCOS, obesity traits, testosterone, and SHBG. We validated our MR results using several different methods, including CAUSE [24], mode-based estimation (MBE), MR-Egger, and the inverse-variance weighted (IVW) method. We did not test the causal relationship between PCOS and testosterone, as testosterone levels are often used to define PCOS. These MR results are shown in the supplementary file 4.

All MR methods suggest that higher BMI could lead to PCOS (effect_CAUSE_ = 0.59, p_CAUSE_ = 6.4*×*10^*−*4^). Most methods suggest that WHR has a causal effect on PCOS, and a few methods suggest that WHRadjBMI is causal. Most of the tests also suggest that low SHBG levels have a significant causal effect on PCOS with effect size about -0.25 (p_CAUSE_ = 0.041). PCOS did not show statistically significant causal effects on any of the tested traits (figure 5), which is consistent with previous MR studies [15]. All MR methods suggest that higher BMI, WHR, and WHRadjBMI may lower SHBG levels and that higher WHRadjBMI may decrease testosterone levels (see supplementary file 4).

**Figure 5:**
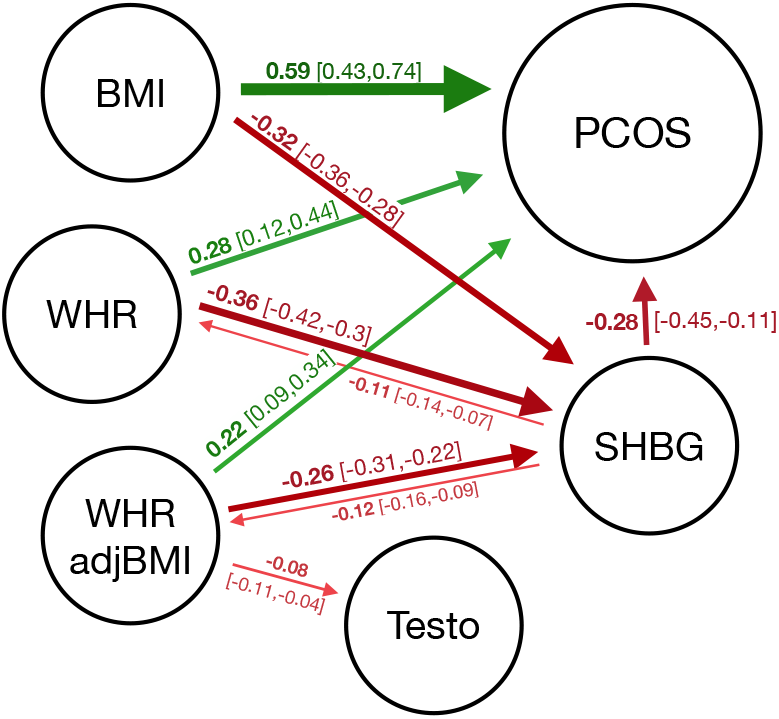
Mendelian randomization estimated causal effects and 95% credible intervals between BMI, WHR, WHRadjBMI, SHBG, Testosterone, and PCOS. Green arrows indicate positive causal effects and red arrows indicate negative causal effects. The thickness and shade of the arrows is proportional to effect size. All estimates are based on the CAUSE model and are significant (p*<*0.05) for the hypothesis that a causal model is a better fit than a sharing model.

#### 2.2.4 Causal relationships between inflammatory biomarkers and PCOS, SHBG, and testosterone

Next we conducted bi-directional MR between PCOS, testosterone, SHBG and the panel of 138 inflammatory biomarkers. For inflammatory markers that are significantly related to any of these traits, we further conducted bi-directional MR between them with BMI and WHR to examine the role of obesity. Our main results were estimated with the MBE MR method, because it has less bias and lower type-I error rates than IVW and is significantly faster than CAUSE. We also examined results from other MR methods to gauge the robustness of the potential associations. Results of MR with inflammatory markers are detailed in supplementary file 5. We did not find statistically significant causality between PCOS and any inflammatory markers based on MR analysis, likely due to the low heritability of the PCOS GWAS.

Twenty-eight inflammatory biomarkers showed a significantly causal effect for SHBG. Of these 28 biomarkers, only CD36antg and CRP were found to decrease SHBG while the other 26 were found to increase SHBG. TWEAK has the strongest causal association with SHBG, an effect that was replicated in all of the MR methods (beta_MBE_ = 0.46, p_MBE_ = 2.46 *∗* 10^*−*219^, beta_IVW_ = 0.260, p_IVW_ = 5.74 *∗* 10^*−*3^, beta_MR-Egger_ = 0.351, p_MR-Egger_ = 1.46 *∗* 10^*−*2^).

Higher genetically-predicted testosterone was found to cause increased levels of CRP (beta_MBE_ = 0.0926, p_MBE_ = 6.91 *∗* 10^*−*7^) (see supplementary table 3). Four inflammatory biomarkers showed significant causal relations with testosterone (FDR_MBE_ =*<* 0.01): genetically-predicted TWEAK (beta_MBE_ = 0.057, p_MBE_ = 3.7 *∗* 10^*−*8^) and MMP9 (beta_MBE_ = 0.043, p_MBE_ = 2.9 *∗* 10^*−*4^) were found to increase testosterone, while genetically-predicted IL2Rb (beta_MBE_ = *−*0.057, p_MBE_ = 7.44 *∗* 10^*−*5^) and IP10 (beta_MBE_ = *−*0.126, p_MBE_ = 1.33 *∗* 10^*−*5^) were found to decrease testosterone. Interestingly, TWEAK is also the only biomarker which shows a causal effect for both testosterone and SHBG.

We next assessed whether inflammation could be partially mediating the causal effect from obesity to sex hormones. BMI showed a significant causal effect on 4 out of 28 biomarkers that had significant causality for SHBG, as visualized in figure 6a. MR suggests that higher BMI may elevate levels of MMP17, IL10Rb, and IL8, which may in turn increase SHBG levels. Higher BMI and testosterone could increase CRP levels, which may lower levels of SHBG.

**Figure 6:**
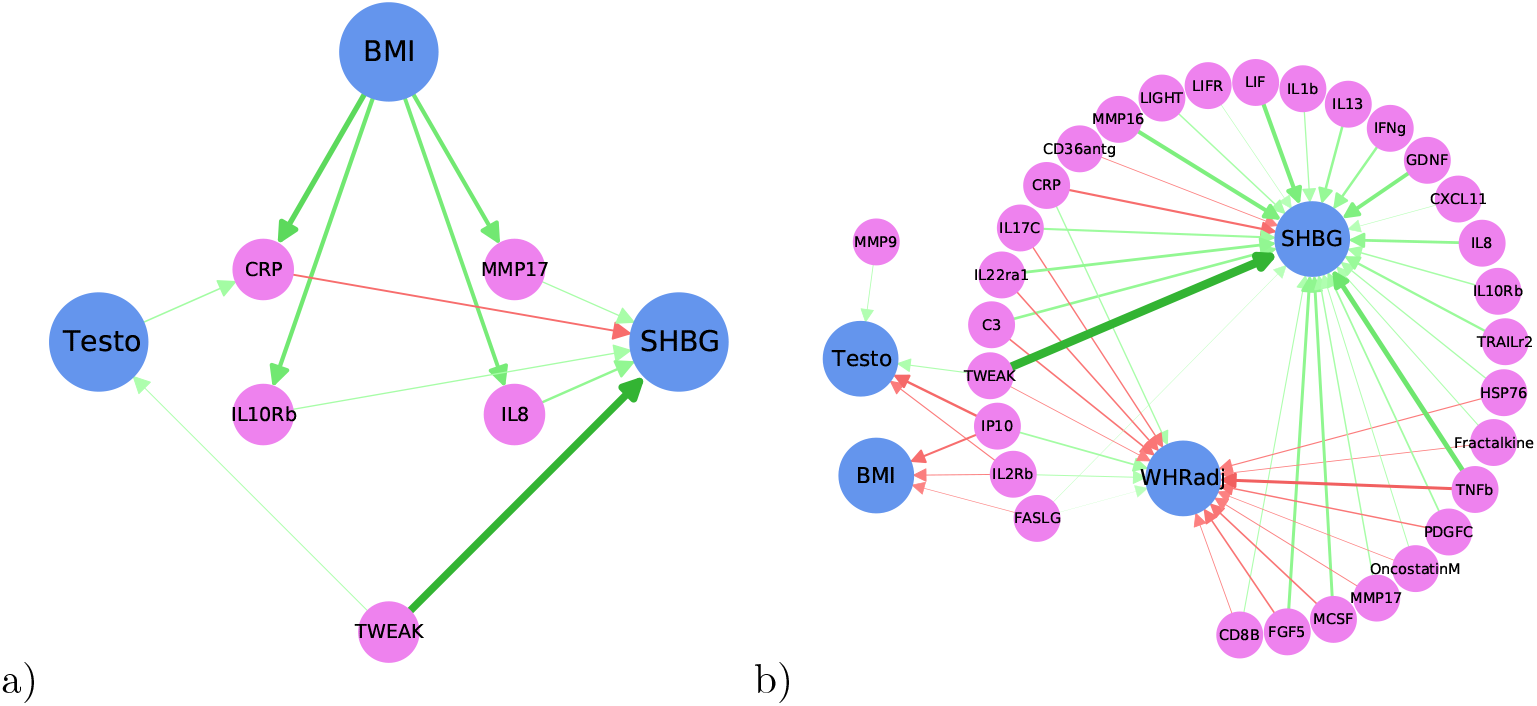
Mendelian randomization results between inflammatory biomarkers and PCOS-related traits. (a) Inflammatory biomarkers may mediate some of the causal relationship between obesity and hormones. (b) Inflammatory biomarkers which significantly regulate SHBG or Testosterone, and the causal relationship of these markers on BMI and WHR adjusted by BMI. Pink circles represent inflammatory markers, blue circles represent traits, and arrows show the direction of causality. Only MBE results are shown in this figure, and only effects with FDR *<* 0.01 are included. Green arrows indicate positive causality, red arrows indicate negative causality, and the width/shade of the arrow indicates strength. Full MR results can be found in the supplementary file 5.

Many of the inflammation markers that were found to increase SHBG were also found to decrease WHRadjBMI (figure 6b), suggesting that these markers could have a protective role. Of the 28 inflammatory biomarkers showing causal effect for SHBG, 15 are also significant for WHRadjBMI, 15 significant for WHR, and 1 (FASLG) for BMI. Details of the MR results are available in the supplementary file 5.

### 2.3 Polygenic risk scores combining causal risk factors improve PCOS prediction

Finally, we created a model for predicting genetic risk of developing PCOS based on the combined polygenic risk scores (PRSs) of PCOS, BMI, WHR, WHRadjBMI, SHBG, and testosterone.

We created PRS from the PCOS, BMI, WHR, WHRadjBMI, SHBG, testosterone summary statistics and applied them to all women in the UK Biobank and Mass General Brigham (MGB) biobank, standardizing each PRS to mean 0 and variance 1. We then created lasso logistic regression models to predict PCOS cases using the PRSs, and compared the prediction to that based solely on PCOS PRS. We trained the logistic regressions and selected hyperparameters via a nested 10-fold cross validation to avoid overfitting.

Our new PRS-based model improved PCOS prediction in both the UK Biobank and the MGB Biobank on held-out data. In the UK Biobank, the area under the ROC curve (AUROC) improved from 0.59 when only using the PCOS PRS to 0.72 when combining the PRSs. In the MGB biobank, the AUROC improved from 0.59 to 0.61. UK Biobank and MGB Biobank model performance and coefficients are shown in figure 7.

**Figure 7:**
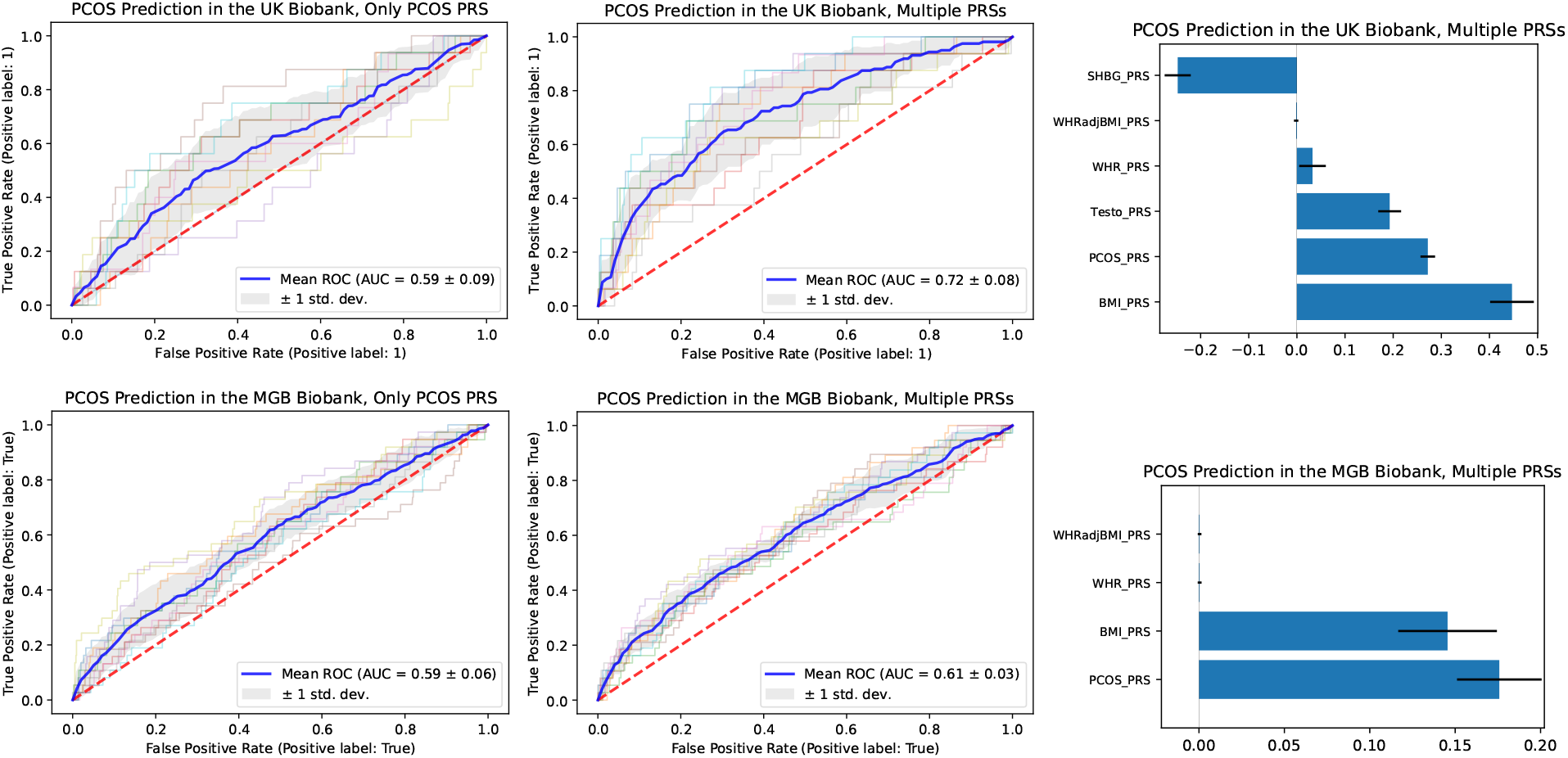
Genetic prediction of PCOS cases in the UK Biobank (top) and MGB biobank (bottom). A nested 10-fold cross validation lasso logistic model was trained on each biobank. The left plots show the receiver operating characteristic (ROC) curve for predicting PCOS cases when only using the PCOS polygenic risk score (PRS). The middle plots show the ROC curve when combining the PCOS PRS with the BMI, WHR, WHRadjBMI, SHBG, and testosterone PRSs, and the left plots show the corresponding model coefficients for each PRS.

The coefficients used to predict PCOS varied between the UK biobank and the MGB biobank. In the UK Biobank, BMI PRS had the largest effect size for predicting PCOS cases, followed the PCOS, SHBG, and the testosterone PRSs. In the MGB Biobank, the PCOS and BMI PRS accounted for most of the prediction power.

Further inclusion of including inflammation PRSs did not improve PCOS predictions in UK biobank or the MGB biobank.

## 3 Discussion

This study investigated genetic correlations and causal relationships between PCOS and obesity, testosterone, SHBG, and biomarkers indicative of a wide-range of inflammatory pathways. Our large GWAS meta-analysis of PCOS identified four novel loci. We demonstrated genetic correlations and potential causal relationships between obesity, testosterone, and SHBG with PCOS, and the potential role of chronic inflammation in these relationships. Interestingly, the genetic correlation between PCOS and testosterone is smaller compared with those with SHBG, BMI, WHR, and WHRadjBMI, despite hyperandrogenism often being used to characterize PCOS. Finally, we showed that incorporating genetics of causal risk markers of PCOS, namely obesity, testosterone, and SHBG, improved genetic risk prediction of PCOS. This study provides evidence that PCOS shares genetic architecture with a range of inflammatory biomarkers. We found significant genetic correlations between PCOS and eleven inflammatory biomarkers, including biomarkers that have not been previously related to PCOS in the clinical literature, such as TRAILr2 (also known as death receptor 5). A previous study has found that TRAILr2 mediates testosterone-driven apoptosis of PCOS granulosa cells in culture [25], which supports the hypothesis that TRAILr2 is involved in the parthenogenesis of PCOS. While our study provides additional evidence that TRAILr2 may be related to PCOS, further research is needed.

Using Mendelian randomization, we found that obesity is likely a causal risk factor for PCOS, while SHBG may protect against PCOS, consistent with previous MR findings [26].

We found through MR that BMI, WHR, and WHRadjBMI all decrease SHBG, suggesting that the previously reported association between obesity and SHBG may be causal [27]. We further found that the relationship between WHR, WHRadjBMI and SHBG is bidirectional, with high SHBG in turn decreasing WHR and WHRadjBMI. This could indicate a positive feedback loop between central obesity and low SHBG, although verification is needed. Consistently, experiments in mice have previously reported that increasing SHBG downregulates de novo lipogenesis and reduces liver fat [28]. Using MR analysis, we found evidence that a broad array of inflammatory biomarkers may affect levels of SHBG, testosterone, and obesity. Of note, higher genetically-predicted TWEAK showed particularly strong causal effects on increasing SHBG and testosterone levels and lowering WHRadjBMI. We further found that testosterone may lead to higher CRP levels, consistent with previous findings that women with PCOS have higher CRP levels than BMI and age-matched controls [18]. These findings support the existing hypothesis that chronic inflammation may lead to dysregulated hormonal production in the ovaries, and also indicate that a more complex profile of inflammatory biomarkers may be involved in this process than previously considered.

The potential causal relationship between many inflammatory biomarkers and SHBG is surprising and requires further explanation. The majority of these inflammatory biomarkers appear to show protective effect by showing a casual effect for increased SHBG and decreased WHRadjBMI and BMI. It should be noted that many of the directions from MR analysis are opposite to the directions of genetic correlations from LDSC. Further study is needed to validate our findings and elaborate on the mechanisms that underlie these results.

While the results of MR suggest causal links, there could also be mediators such as insulin resistance, diabetes, CVD, metabolic syndrome, and other conditions that are causal for testosterone, SHBG levels, and chronic inflammation. For this reason it is important to interpret the results as one piece of evidence and apply general caution as needed whenever dealing with Mendelian randomization studies.

Taking advantage of these related traits, we created a model to improve genetic prediction of PCOS by incorporating multiple PRS scores, and found that including information from the genetics of obesity, SHBG, and testosterone significantly improved PCOS prediction in two independent biobanks. This result indicates that the power of current PCOS GWAS is still limited for polygenic risk prediction and can be improved by incorporating PRSs from genetically-related traits. Since PCOS is hard to diagnose, up to 75% of cases remain unidentified [29], and using genetics to flag potential cases could help improve detection of high risk population and enable early intervention.

It is worth noting, though, that even the improved AUROC remains relatively low—0.72 in the UK Biobank and 0.61 in the MGB Biobank. A likely reason for this is that there are probably many people with undiagnosed or unreported PCOS in both biobanks. The population prevalence of PCOS is predicted to be between 5% and 15% in European populations [30], but its reported prevalence in the UK Biobank is approximately 0.01% and in the MGB biobank approximately 4.7%. PCOS is challenging to properly diagnose, requiring multiple clinical and laboratory assessments including a pelvic ultrasound. It is possible that many of the “false positives” in the model could be women who have PCOS but are undiagnosed. Another possibility is that these polygenic risk scores are not accurate enough to effectively separate women with PCOS from those without.

When we added inflammatory biomarkers into the model for genetic prediction of PCOS, they did not further improve PCOS prediction in either biobank. It is possible that any effect of these biomarkers on obesity, testosterone, and SHBG is already captured by PRSs of obesity, testosterone, and SHBG.

Our study has several limitations. The aforementioned under-diagnosis of PCOS cases not only affects the ability to create PRS scores within the biobanks, but also the power of PCOS GWAS due to false negatives within the biobanks. Furthermore, MR analysis has limitations and any violations to the assumptions may bias the results—MR can only suggest potential causal relationships that must be verified in clinical studies. Importantly, this study was limited to individuals of European ancestry due to the small case counts in biobanks; PCOS has high prevalence globally and thus future studies including diverse populations are needed [31, 32]. Since PCOS may present itself differently between populations, it is especially important that biobanks increase diversity to improve PCOS research for all.

In summary, our study identifies four novel genetic loci for PCOS and demonstrates shared genetic architecture and potential causal relationships between PCOS and obesity, SHBG, testosterone, and inflammation. Together, these results support theories that immune responses are altered in PCOS patients and that chronic inflammation plays a role in dysregulating testosterone and SHBG levels.

## 4 Methods

### 4.1 Data Sources

#### 4.1.1 PCOS Summary Statistics Data

We obtained PCOS summary statistics from the PCOS GWAS in the FinnGen and Estonia Biobanks [9] and a recent cross-population PCOS meta-analysis in European populations [10]. In the FinnGen and Estonia biobanks there were a total of 3,609 cases and 229,788 controls. All cases were self-reported and all other women were considered controls. In Tyrmi et al. [9], GWASs were conducted on the FinnGen and Estonia cohort before they were combined in an inverse-variance-weighted meta-analysis. Both GWASs used population-specific imputation panels: the Sequencing Initiative Suomi V3 [33] for FinnGen and Mitt et al. [34] for EstBB. Associations were run using the SAIGE generalized mixed model [35], and included age, genotype batches, and PCs 1-10 as covariates. Due to including rarer variant alleles, Tyrmi et al. [9] includes 22.8 million SNPs.

In Day et al. [10] there are 4,137 PCOS cases and 20,129 controls pooled together in a fixed-effect, inverse-weighted-variance meta-analysis from 6 cohorts (Rotterdam, British Birth Cohort, Estonian Genome Center of the University of Tartu (EGCUT), deCODE genetics, Chicago, and Boston). In total there are 8.8 million SNPs. The Estonian cohort used in Day et al. [10] has 157 cases and 2807 controls which overlap with cases and controls used in Tyrmi et al. [9]. This causes a 2% overlap between the two studies. Since we did not observe any inflation in LD regression intercept and the overlap is small, we assume this will not create disproportionate effects and continue with the analysis.

#### 4.1.2 Obesity Summary Statistics Data

We obtained BMI summary statistics data from Yengo et al. [36], a meta-analysis of GIANT and UKB for a total N of 681,275 men and women. WHR and WHRadjBMI summary statistics are from a GIANT and UKB meta-analysis, conducted by Pulit et al. [37]. We used female-specific summary statistics, resulting in N=263,148 for WHR and N=262,759 for WHRadjBMI.

#### 4.1.3 Testosterone and SHBG GWAS

SHBG and testosterone serum level GWAS were conducted using the UK Biobank females who identified as “White British” and matched ancestry based on principal components. Serum levels for SHBG were available for 190,366 women and serum levels for total testosterone were available for 176,687 women. GWAS was conducted using the BOLT-LMM algorithm, adjusting for the first 20 PCs, age, age^2^, menopausal status, pre-menopausal oral contraceptive use, and postmenopausal hormone therapy use [38]. We replicated all genetic correlation analysis with pre-menopausal testosterone and SHBG and found the same results, so kept the analysis using the joined pre and post-menopausal serum levels in order to increase power.

#### 4.1.4 PCOS in the UK Biobank

Samples in the UK Biobank were used to train and test the polygenic risk score model for improved genetic prediction of PCOS. We used UK Biobank release version 3 with participants limited to females who self-identified as “White British” and matched ancestry based on principal components. PCOS cases were defined by self report, by which there are 159 cases.

Controls were filtered based on Rotterdam phenotypes to try to minimize the number of false negatives. Controls were selected from females that did not report ICD codes indicating excess androgen or irregular menstruation. ICD codes used to indicate excess androgen and irregular menstruation are the same as in Zhang et al. [39]. To enable a more balanced ratio for classification in the logistic regression, nineteen controls were randomly matched by age to each case to match the lower estimated population prevalence of 5%.

#### 4.1.5 PCOS in the MGB Biobank

Samples, genomic data, and health information were obtained from the Mass General Brigham Biobank, a biorepository of consented patients samples at Mass General Brigham (parent organization of Massachusetts General Hospital and Brigham and Women’s Hospital). These samples were also used to train and test the polygenic risk score model for improved genetic prediction of PCOS. Participants were limited to females who self-identify as white.

PCOS cases were defined by ICD self report, through which there are 374 cases. The control criteria in the MGB biobank was the same as in the UK biobank. Women who reported ICD codes indicating excess androgen or irregular menstruation were removed from the analysis to minimize false negatives. Using this criteria, there were a total of 7,553 controls.

#### 4.1.6 Inflammatory Biomarker Data and Summary Statistics

To characterize inflammation on both cellular and molecular level and from multiple inflammatory pathways, we curated GWAS for a total of 138 inflammatory biomarkers.

##### Blood immune cell types

GWAS for 6 biomarkers are curated for counts of white blood cells, neutrophils, lymphocytes, monocytes, eosinophils, basophils. We conducted GWAS for these cell types (inverse normal transformed) in the UK Biobank using BOLT-LMM, adjusting for 20 PCs, age, age2, sex, and study center. The GWAS summary statistics were further combined with published summary statistics from the Biobank Japan (BBJ) [40] using an inverse-variance weighted meta-analysis using METAL. **Lymphocyte subtypes:** GWAS for 6 lymphocyte subsets, including CD4+ T cells, CD8+ T cells, CD56+ natural killer (NK) cells, CD3+ T cells, CD19+ B cells, and the derived measure CD4:CD8 ratio, are obtained from Ferreira et al. [41].

##### Molecular biomarkers of inflammation

GWAS for 126 biomarkers, including CRP and biomarkers indicative of diverse inflammatory pathways, were curated by meta-analysis of published and newly conducted GWAS. For CRP, GWAS were conducted in the UK Biobank (similar methods with those for immune cell subtypes); the NHS/HPFS (using Rvtests [42], after inverse-normal transformation, adjusting for the first 10 PCs, age, sex, cohorts and sub-studies); and GWAS summary statistics from UKB and NHS/HPFS were further meta-analyzed with published GWAS summary statistics from the Biobank Japan. For other biomarkers, we conducted GWAS for adiponectin, leptin, ICAM1, IL6, TNFaR1, TNFaR2, and IL18 in the NHS/HPFS; we acquired other biomarkers GWAS summary data from several publicly available sources including the Ahola-Olli et al. [43], Dastani et al. [44], and Kilpeläinen et al. [45], and GWAS of proteomics including Suhre et al. [46], Sun et al. [47], and Ferkingstad et al. [48]. For GWAS of proteomics measured using aptamer based SOMAscan platform, some markers in the proteomics dataset had different aptamers for the same protein target; we chose the GWAS for the aptamer with more genome-wide significant signals. We conducted meta-analysis for the same circulating protein biomarkers using the METAL [49] with the inverse-variance-weighted method. The detailed information on the meta-analysis of these inflammatory biomarkers is included in the supplement.

### 4.2 Analyses

#### 4.2.1 PCOS Meta-Analysis and Genome-Wide Significant Loci

We combined Tyrmi et al. [9] summary statistics and Day et al. [10] in METAL [49] using the inversevariance-weighted method. The summary statistics were in genome build GRCh37 and analyzed in PLINK [50] to clump loci using the setting *p*_1_ = 5*e −* 8, *p*_2_ = 1*e −* 5, clump-kb = 1000, and *r*2 = 0.01. We compared clumps that reached genome-wide significance in the meta-analysis to PCOS-associated SNPs from previous studies in order to identify novel loci. We consulted GWAS catalog to check previous studies for significant loci [51] and used locuszoom to visualize the GWAS [52].

#### 4.2.2 Genetic Correlations using LDSC

We ran LDSC [53] to find the genetic correlations. We first found genetic correlations between PCOS, BMI, WHR, WHRadjBMI, SHBG, and testosterone, and later we calculated genetic correlations between these traits and each of the 138 inflammation markers. We used a SNP list from HapMap3 [54], computed LD scores in European ancestry from 1000 Genomes [55], and limited SNPs to those with MAF*>*0.01.

During the inflammation analysis, p-values were corrected via false discovery rate (FDR), and only correlations with FDR*<* 0.05 (138 inflammation markers *×* 6 traits = 828 tests) were included in the analysis.

#### 4.2.3 Mendelian Randomization

We conducted Mendelian randomization to test causal relationships with PCOS and related metabolic, hormonal, and inflammatory traits. We tested for causality in both directions between PCOS, each obesity trait, each hormonal trait, and each inflammation marker. Since androgen excess can be used as part of diagnosing PCOS, testosterone and PCOS break some of the MR assumptions, and thus their causality was not tested.

For trait-to-trait MR analyses, we implemented several MR models. First we used the Causal Analysis Using Summary Effect estimates (CAUSE) model [24]. CAUSE accounts for correlated and uncorrelated horizontal pleiotropic effects and thereby avoids more false positives. To find significant SNPs that are not in LD, we used PLINK [50] with parameters *p*1 = 5*e −* 8, *p*2 = 5*e −* 8, clump-kb = 1000, and *r*2 = 0.01. We also compared the CAUSE estimates to estimates calculated via the mode-based estimate (MBE) [56], MR-EGGER [57], and inverse variance weighting (IVW) methods. We calculated each of these tests via the Mendelian randomization R package [58].

For trait-to-inflammation or inflammation-to-trait MR analyses, we only used the Mendelian randomization R package [58]. For every test we mainly used mode-based estimate (MBE) method, which allows relaxation of the instrumental variable assumptions and has less bias and lower type-I error rates than other methods [56]. We also looked for consistency using the IVW, and MR-EGGER methods. The CAUSE method was not implemented, since it takes a too long of a time to run each test. We only report as significant associations with FDR *<* 0.01 (138 tests) to minimize the number of false positive correlations.

#### 4.2.4 Polygenic Risk Score Model to Improve PCOS Prediction

Our goal was to improve PCOS prediction by combining the PCOS polygenic risk score (PRS) with genetic scores of related risk factors. First we included the PRSs of PCOS, obesity measurements, SHBG, and testosterone, and we later added 138 inflammatory PRSs to further improve the prediction. We compared these models to a model that only considers the PCOS PRS.

To calculate the PRSs, we used sBayesR [59] to create a list of variants and effect sizes for each trait (PCOS, Testosterone, SHBG, BMI, WHR, WHRadjBMI). We used decreasing p-values starting at 0.5, using the highest possible p-value where sBayesR converged. For the inflammation biomarkers, we used PRS-CS with the 1000 Genomes Phase 3 reference panel [55] and the PRS-CS auto setting to create a list of variants and effect sizes [59]. Then we used PLINK [50] to apply these SNP effects to create polygenic risk scores for every female of European descent in the UK Biobank and MGB Biobank.

We created lasso logistic regression models to predict PCOS cases from multiple PRSs in both the MGB biobank and UKB biobank. The logistic regressions were trained via a nested cross validation (CV), with a 10-fold outer CV and 5-fold inner CV. For the outer loop, the whole dataset was randomly split into 10 equal groups. Each group was used once as a holdout set, with the remaining 9 groups used as the training set; a randomly-split inner 5-fold cross validation within only the training set was used to tune the regularization parameters; the resulting regression model was used to predict the out-of-sample 10th group. The prediction performance was evaluated using the area under the receiver operating characteristic (ROC) curve. In evaluation, PCOS cases were weighted higher than controls (weights were inversely proportional to class frequencies) to account for class imbalances. Scikit-Learn was used to implement all models [60].

## Data Availability

All data produced in the present study are available upon reasonable request to the authors

## 5 Acknowledgments

L.K.P. gratefully acknowledges the support of the U.S. Department of Energy (DOE) through the Los Alamos National Laboratory (LANL) LDRD Program and the Center for Nonlinear Studies for this work. All authors thank Jocelyn Neri for help implementing Rotterdam Criteria in the MGB biobank. This research was conducted using the UK Biobank Resource under Application #45052. The authors would like to thank all participants and staff from the Mass General Brigham Biobank and UK Biobank. We further thank the Channing Network Health Division and all participants and staff in the Nurses Health Study.

## 6 Author Contributions

L.K.P., G.B., and L.L. designed the research study. L.K.P and G.B. lead all analyses. J.L., X.H., L.P. and G.B. curated the meta analyses of inflammatory biomarkers. J.H. conducted the GWASs for SHBG and testosterone in the UKB. Z.H. helped create polygenic risk scores. T.P. and S.M. provided guidance on clinical PCOS phenotypes and diagnosis, A.Y.M. provided guidance on inflammatory marker interpretation, and J.T. provided guidance on GWAS hit interpretation. L.K.P. and G.B. wrote the paper, with input from all authors. All authors provided valuable feedback and helped shape the analysis and manuscript.

### 7 Funding Source

The Nurses’ Health Study, Nurses’ Health Study II, and Health Professionals Follow-up Study are supported by NIH grants UM1CA186107, R01CA49449, U01CA176726, R01CA67262, and U01CA167552.

